# Independent morphological variables correlate with Aging, Mild Cognitive Impairment, and Alzheimer’s Disease

**DOI:** 10.1101/2022.01.10.22268812

**Authors:** Fernanda Hansen Pacheco de Moraes, Felipe Sudo, Marina Carneiro Monteiro, Bruno R. P. de Melo, Paulo Mattos, Bruno Mota, Fernanda Tovar-Moll

**Affiliations:** Brain Connectivity Unit, Instituto D’Or de Pesquisa e Ensino (IDOR), Rua Diniz Cordeiro 30, Botafogo, Rio de Janeiro 22281100, RJ, Brazil; Memory Clinic, Instituto D’Or de Pesquisa e Ensino (IDOR), Rua Diniz Cordeiro 30, Botafogo, Rio de Janeiro 22281100, RJ, Brazil; Instituto de Física - Universidade Federal do Rio de Janeiro (UFRJ), Av. Athos da Silveira Ramos, 149 Centro de Tecnologia - bloco A - Cidade Universitária, Rio de Janeiro, 21941-972, Brazil

**Keywords:** Cortical Folding, Aging, Alzheimer’s Disease, Mild Cognitive Impairment

## Abstract

This manuscript presents a study with recruited volunteers that comprehends three sorts of events present in Alzheimer’s Disease (AD) evolution (structural, biochemical, and cognitive) to propose an update in neurodegeneration biomarkers for AD. The novel variables, K, I, and S, suggested based on physics properties and empirical evidence, are defined by power-law relations between cortical thickness, exposed and total area, and natural descriptors of brain morphology. Our central hypothesis is that variable K, almost constant in healthy human subjects, is a better discriminator of a diseased brain than the current morphological biomarker, Cortical Thickness, due to its aggregated information. We extracted morphological features from 3T MRI T1w images of 123 elderly subjects: 77 Healthy Cognitive Unimpaired Controls (CTL), 33 Mild Cognitive Impairment (MCI) patients, and 13 Alzheimer’s Disease (AD) patients. Moreover, Cerebrospinal Fluid (CSF) biomarkers and clinical data scores were correlated with K, intending to characterize health and disease in the cortex with morphological criteria and cognitive-behavioral profiles. K distinguishes Alzheimer’s Disease, Mild Cognitive Impairment, and Healthy Cognitive Unimpaired Controls globally and locally with reasonable accuracy (CTL-AD, 0.82; CTL-MCI, 0.58). Correlations were found between global and local K associated with clinical behavioral data (executive function and memory assessments) and CSF biomarkers (t-Tau, A*β*-40, and A*β*-42). The results suggest that the cortical folding component, K, is a premature discriminator of healthy aging, Mild Cognitive Impairment, and Alzheimer’s Disease, with significant differences within diagnostics. Despite the non-concomitant events, we found correlations between brain structural degeneration (K), cognitive tasks, and biochemical markers.

## Introduction

Alzheimer’s Disease (AD) is the most common dementia worldwide. Besides being widely studied, there is a lack of knowledge on global brain mechanics, reflected in its morphology during the disease stages. The clinical diagnosis of Alzheimer’s Disease relies on episodic memory impairment, neuropsychological assessment, at least one abnormal biomarker among Cerebrospinal fluid (CSF) analysis, and neuroimaging (PET and MRI) (1). There is some cognitive dysfunction in its preclinical or prodromal stage, Mild Cognitive Impairment (MCI), but minor extension as in dementia. MCI patients can be diagnosed as amnestic or anamnestic, depending on memory loss presence, and by the decline of single or multiple domains (2). AD is also characterized by the concentration of A*β*1-40, A*β*1-42, and total Tau protein on the CSF, which is correlated with findings of amyloid plaques and Tau tangles on histopathological examinations (3, 4). In addition, new biomarkers for AD have been suggested based on the pathology’s inflammation, as Lipoxin, which regulates chronic inflammatory processes resolution (5). In structural images, Alzheimer’s Disease is characterized by brain atrophy, which includes volume reductions in the medial temporal lobe and hippocampus, grey matter loss, and reduced Cortical Thickness (6), currently used as biomarkers (7, 8).

However, computing brain folds (9), arising from the cortical folding (or gyrification) process is an upcoming morphological measurement of the brain that is related to age (10– 12) and neurological pathologies (Major Depressive Disorder, Bipolar Disorder, Schizophrenia (13), Anorexia Nervosa (14), Autism (15), and Alzheimer’s Disease (16)). Cortical folding can be measured from a structural T1-weighted MRI: (i) by using the primary parameter of cortical folding, the Gyrification Index (GI), that is the ratio of the Total Area (A_T_) of the brain and its Exposed Area (A_E_) (17), (ii) by calculating the fractal dimension (18) or (iii) by calculating an index (k) derived from a mathematical relation of the, A_T_, A_E_), and the Average Thickness (T) (19). The latter was proposed as a physics-based model for cortical folding by Mota & Herculano-Houzel, predicting a power-law relationship between Cortical Thickness, Exposed, and Total Areas experimentally confirmed on the brains of 55 mammals, including humans. Its linear coefficient, k, is a natural variable to describe brain morphology, while the angular coefficient *α* is the fractal index and universal constant, with theoretical value 1.25 and calculated as 1.305 (19).

A follow-up study by Wang et al., 2016 (20) described the universal law of gyrification for the human brain across gender, age, and pathology, with further confirmation that the universal law is also respected in smaller fractions of the cortex by applying the at four lobes (Frontal, Occipital, Parietal, and Temporal Lobes)(21). In Wang et al. 2021 (22), by taking the logarithmic variables log_10_A_T_, log_10_A_E_ and log_10_ T^2^ as the base space, it was proposed a new set of independent variables to described brain morphology derived from perpendicular planes of log_10_ k: K (Equation 1), representing the axonal tension, S (Equation 2), that encapsulates brain shape, and I (Equation 3), representing brain volume. Combined, those three variables could help distinguish pathological events similar to age effects, such as AD.

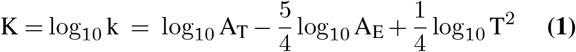

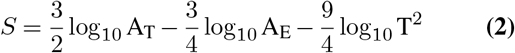

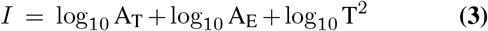

As a practical application of Mota, Herculano-Houzel, and Wang’s cited works, we propose an improvement in the differential diagnosis of AD, MCI, and CTL, with cortical folding independent variables. Therefore, we suggest that AD effects in brain morphology are similar to accelerated aging in the K, I, and S base space.

Further, we correlate those variables, representing pathological structural changes, with neuropsychological tests used to diagnose dementia (cognitive function, working and episodic memory, and memory estimation), CSF biomarkers related to Alzheimer’s Disease (total Tau, A*β*1-40, A*β*1-42), and Lipoxin, a regulator of chronic inflammatory processes resolution(5).

## Results

With 123 subjects (77 CTL, 33 MCI, and 13 AD) (Demographics and summary for each group in Supplementary Notes, Table 3), the data fits the model with *α* = 1.13 ± 0.03 (95% CI: 1.07; 1.19), statically different from the theoretical value, 1.25 (Student’s t = 3.65, p = 0.00032). Healthy aging reduces brain gyrification in terms of *α* (Pearson’s r = -0.79, p = 0.0345) (Supplementary Note 2), K (Pearson’s r = -0.32, p <0.0001) (Figure 1), and I (Pearson’s r = -0.47, p <0.0001).

**Fig. 1.**
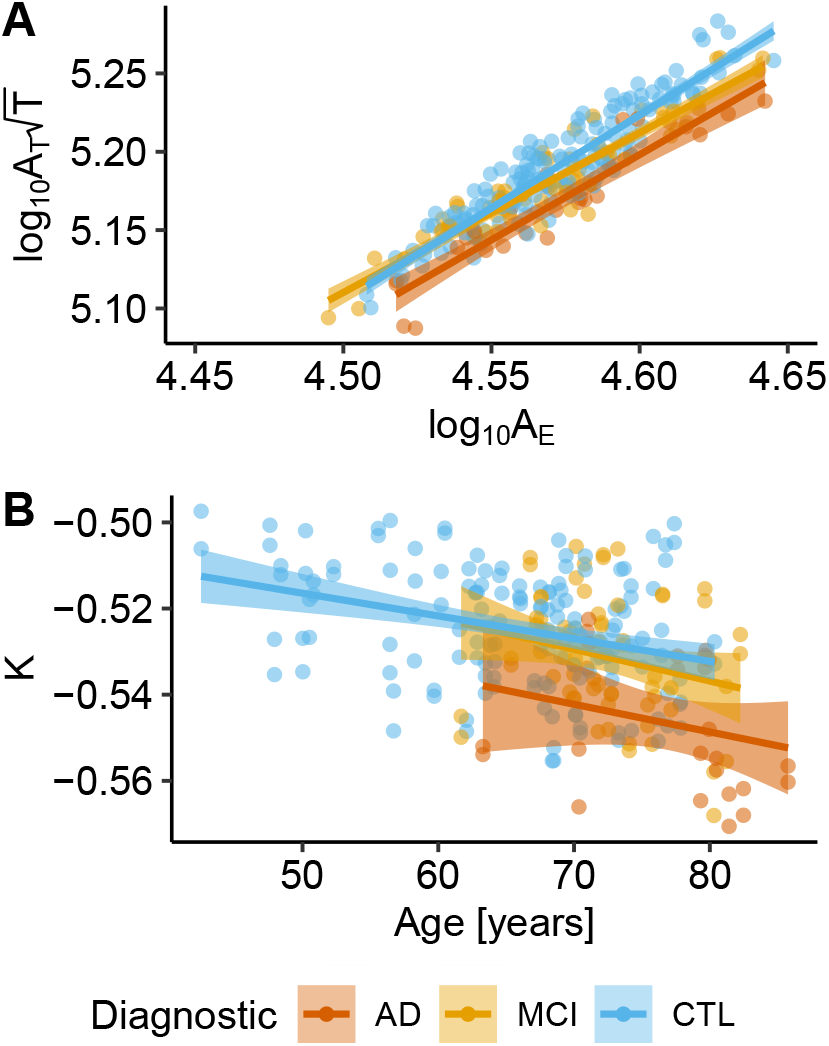
Age and diagnostic effects in cortical gyrification. (A) Linear fitting with 95% Confidence Interval (CI) for the model variables in each Diagnostic group, CTL (adjusted R^2^ = 0.85, p < 0.0001), MCI (adjusted R^2^ = 0.88, p < 0.0001), and AD (adjusted R^2^ = 0.86, p < 0.0001). As the severity of the disease increase, the linear tendency is downshifted, with smaller linear intercepts (K). (B) K linear tendency across age with 95% CI for the three diagnostics groups: AD (adjusted R^2^ = 0.026, p = 0.21), MCI (adjusted R^2^ = 0.044, p = 0.0051), and CTL (adjusted R^2^ = 0.097, p < 0.0001).

### Diagnostic discrimination and prediction

K is different for the diagnostics groups in the hemisphere analysis (ANOVA F = 28.27, p < 0.0001) (p.adj < 0.01 for all pairwise comparisons), meaning a global structural change with the pathology. The decrease of K with disease presents a similar pattern of the decrease with healthy aging, in which Cortical Thickness, Exposed, and Total Areas are reduced. Taking into consideration specific lobes, AD, MCI, and CTL presented differences in gyrification in all lobes (Frontal lobe, F = 14.71 p <0.0001; Occipital lobe, F = 10.07, p <0.0001; Parietal lobe, F = 16.65, p <0.0001 and Temporal lobe, F = 28.49, p <0.0001). Subsequent pairwise comparisons showed significant differences between CTL-AD and MCI-AD for all lobes, and a significant difference between CTL and MCI for the Temporal lobe (SI Appendix, Fig. S1). There is no statistical power to infer if the difference in K between AD and CTL, increases with age (SI Appendix, Fig. S2).

To compare K and Cortical Thickness (log_10_T) discriminating power, we evaluated their optimal cut-offs in raw data and after age correction (Figure 2). K optimal cut-off for discriminating Alzheimer’s Disease and Cognitive Unimpaired Controls is -0.54, and for discriminating Mild Cognitive Impairment and Controls, -0.53. K has excellent accuracy and reasonable specificity discriminating AD from CTL (ACC = 0.82, specificity = 0.86), and low sensitivity (0.58), while log_10_T (ACC = 0.73) has a balanced trade-off with specificity and sensitivity (0.77 and 0.73 respectively). Discriminating MCI from CTL is challenging for both K (ACC = 0.60) and log_10_T (ACC = 0.55). K (after age correction) cut-points for lobes are described in Table 1 (expanded results in SI Appendix, Table S2), by which it is possible to verify local regions more prone to diagnostic discrimination.

**Fig. 2.**
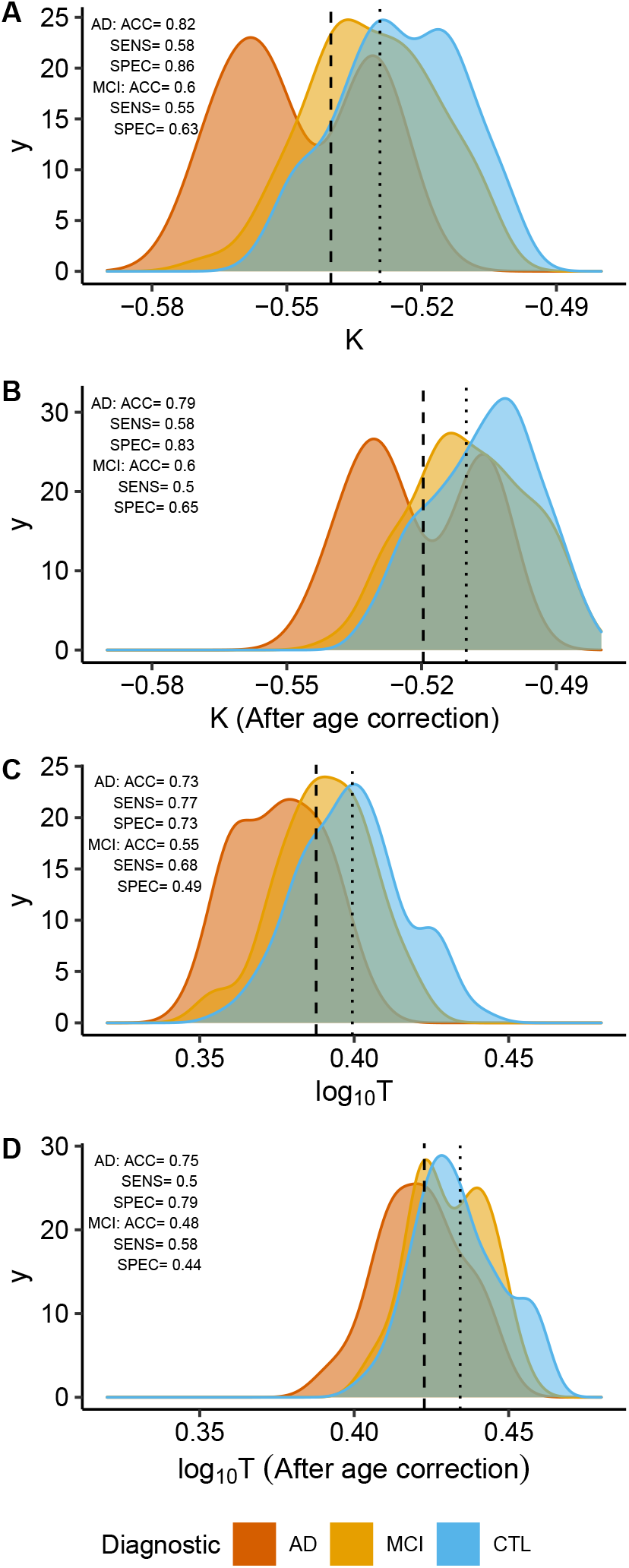
Optimal cut-off (maximum sensitivity + specificity) for K and Cortical Thickness including results with removed age effect (“age correction”). The dashed line represents optimal cut-off to discriminate AD and CTL, and the dotted line represents optimal cut-off to for MCI and CTL. “ACC” - accuracy, “SPEC” - specificity, and “SENS” - sensibility. (A) For K, the optimal cut-off for the CTL-AD contrast is -0.54 and CTL-MCI, -0.53. (B) For K, after age correction, the optimal cut-off for CTL-AD = -0.52 and CTL-MCI = -0.51. (C) For log_10_T, the optimal cut-off for CTL-AD = 0.39 mm and CTL-MCI = 0.40 mm. (D) For log_10_T, after age correction, the optimal cut-off for CTL-AD = 0.43 mm and CTL-MCI = 0.44 mm.

**Table 1.**
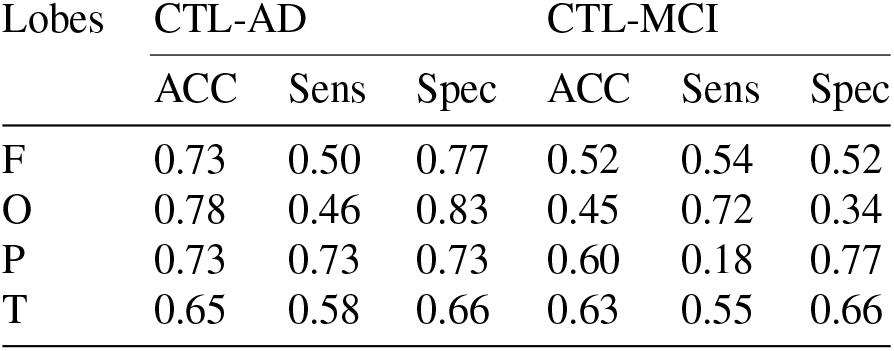
Optimal cut-off (maximum sensitivity + specificity) for K (age-corrected) at each lobe (F - Frontal, O - Occipital, P - Parietal, and T - Temporal lobes.). Cut-off, accuracy, sensibility, and specificity for discriminating pairwise diagnostic groups.

| Lobes | CTL-AD |  |  | CTL-MCI |  |  |
| --- | --- | --- | --- | --- | --- | --- |
|  | ACC | Sens | Spec | ACC | Sens | Spec |
| F | 0.73 | 0.50 | 0.77 | 0.52 | 0.54 | 0.52 |
| O | 0.78 | 0.46 | 0.83 | 0.45 | 0.72 | 0.34 |
| P | 0.73 | 0.73 | 0.73 | 0.60 | 0.18 | 0.77 |
| T | 0.65 | 0.58 | 0.66 | 0.63 | 0.55 | 0.66 |

### Aging and pathological morphology alterations

It is essential to notice that despite aging and AD share similar changes in neurodegenerative-related gyrification patterns, our data suggest that the underlying neurobiological mechanisms related to cortical shrinking are distinct in each group. We compared the hemispheric values of K, S, and I for two groups (66 to 75 years old and 76 to 85 years old) to discriminate the effect of aging or AD pathology on brain morphometry. ANOVA showed significant effect on the Diagnostic:Age group interaction in K (F = 13.3, p < 0.0001), the tension component, the shape component S (F = 4.70, p = 0.000412), and I (F = 5.961, p < 0.0001), the volume component. Further, combined, K, S, and I trajectories suggest that MCI is an intermediary stage between healthy aging and Alzheimer’s Disease. The trajectories suggest that the AD group reaches a plateau across brain tension and shape (K and S), with the most noticeable changes in I, the volume component (Figure 3).

**Fig. 3.**
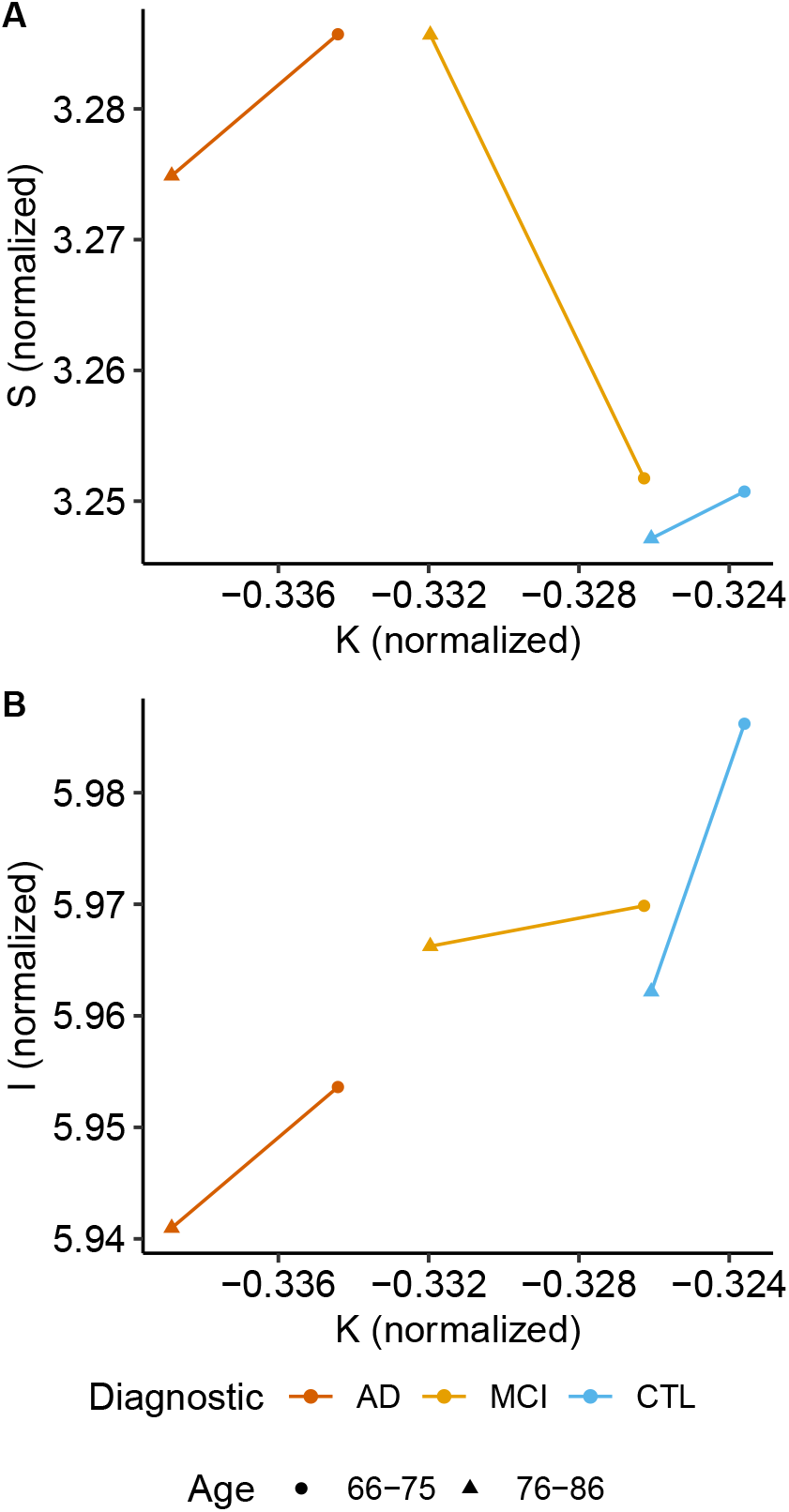
Morphological trajectory traced across the normalized independent components K, S, and I. We normalized the variable to the unity vectors providing comparable scale for the differences in both axes. Groups were divide in two subgroups, subjects with age between 65 and 75 years old (CTL N = 67, MCI N = 24, AD N = 4) and subjects with ages between 76 and 85 years old (CTL N = 10, MCI N = 9, AD N = 9).

### Behavioral and CSF data correlation

Further, we aimed to verify if the AD-related morphological brain changes would be correlated to clinical and behavioral variables and CSF biomarkers of the disease. We found significant correlations between executive function (Cognitive Index, Digit Span Backwards), cognitive flexibility tasks related to memory (TMT B-A), and episodic memory (RAVLT A7/A5) with K and Cortical Thickness (Table 2 and extended results in SI Appendix, Table S3). The severity of cognitive symptoms was associated with decreased gyrification and decreased Cortical Thickness.

**Table 2.** Pearson Correlations (r) for behavioral assessments and morphological parameters, K and log_10_ Cortical Thickness (log_10_T) after age correction. p-value was corrected (Bonferroni) for multiple comparisons within Clinical Assessment and morphological measurement. ROI codes: H - Hemisphere, F - Frontal Lobe, O - Occipital Lobe, P - Parietal Lobe and T - Temporal Lobe.

| Clinical Assessment | ROI | K (r) | $\log_{10}T$ (r) |
| --- | --- | --- | --- |
| Cognitive Index | H | 0.32*** | 0.24*** |
| RAVLT A7/A5 | H | 0.27*** | 0.26*** |
| RAVLT A7/A5 | T | 0.23*** | 0.34*** |
| TMT B-A | H | -0.22** | -0.067 |
| TMT B-A | F | -0.11 | -0.080 |
| Digit Span Backward | H | 0.22** | 0.099 |
| Digit Span Backward | F | 0.17* | 0.074 |
| $A\beta_{1-40}$ | H | -0.054 | -0.18 |
| $A\beta_{1-42}$ | H | 0.21 | -0.012 |
| t-Tau | H | -0.18 | -0.29** |
| $A\beta_{1-42}/A\beta_{1-40}$ | H | 0.15 | 0.14 |
| t-Tau/$A\beta_{1-42}$ | H | -0.22 | -0.16 |
| t-Tau/($A\beta_{1-42}/A\beta_{1-40}$) | H | -0.20 | -0.24* |
| Lipoxin | H | 0.11 | -0.035 |

p-value codes: \*\*\* $\leq 0.001$ , \*\* $\leq 0.01$ , \* $\leq 0.05$

For CSF biomarkers, we found significant correlations between K and the concentration of t-Tau, concentration of A*β*1-42, and ratios of Tau and *Aβ* concentrations. Decreased gyrification index was associated with elevated concentrations of t-Tau and its ratios with *Aβ* in CSF and increased concentrations of A*β*1-42 (Table 2 and extended results in SI Appendix, Table S3).

## Discussion

Structural MRI imaging biomarkers have been largely studied, including gyrification applied to AD (23) and MCI (24). However, investigating cortical morphological measurements (or their combinations) is not a straightforward task since only a few of these parameters will lead to biological interpretations and adequate characterizations of the event in the study. Inspired by the cortical folding model proposed by Mota & Herculano-Houzel (19), we aim to investigate an improvement on structural biomarkers with independent components to discriminate diseased and healthy aged brains better. The hypothesis relies on the singularity of brain gyrification variable K, its biological meaning, and its low variance across species, especially in healthy adult humans. We have shown that the gyrification variable K discriminates patients with AD from MCI and age-matched controls. Further, we have shown that structural damages described by K correlate with cognitive decline and biochemical CSF changes related to AD.

From the chosen independent parameters, K is a natural descriptor of cortical folding and global brain morphology as it is: i) a universal law for mammals, including lissencephalic and cetaceans; ii) based on physical properties, iii) aggregate structural information from Areas and Cortical Thickness, iv) is based on empirical evidence (19) and, v) it is proven to have a robust behavior when applied to smaller regions of interest in the human cortex (21). In addition, brain gyrification variables S and I are mathematically derived from K, but they are independent. Changes on K do not imply changes in S and I, as previously described (22).

Therefore, corroborating the primary hypothesis in this study, our results suggest that K is a sensitive variable for differentiating AD patients, MCI patients, and normal aging subjects, with complex biological and theoretical backgrounds compared to other previously established structural biomarkers such as Cortical Thickness. More specifically, in the Alzheimer’s Disease application, K is an excellent candidate to become a neurodegenerative biomarker in the NIA-AA AT(N) framework (25). Moreover, including S and I to create a morphological trajectory, our findings drive to the hypothesis that morphological degeneration in Alzheimer’s Disease could be interpreted as a premature or accelerated form of cortical aging and unfolding of the brain. In this study, the subject’s assessment covered multiple clinical and behavioral domains and investigated biochemical CSF biomarkers of neurodegeneration, allowing us to confirm that alteration on cortical gyrification (in K) is correlated to changes in cognitive function and biochemical markers in AD pathology.

### Model fitting

To investigate the hypothesis properly, we first verified if the model proposed by Mota & Herculano-Houzel was adequately fitted to our dataset. We compared *α* for each group with the theoretical value, 1.25 (19) to verify our data fits the proposed model for gyrification. *α* was defined as 1.25 to calculate K, S, and I to study how cortical gyrification changes within the selected diagnostics, allowing comparisons with previous investigations.

The presented data fit the model and has a slope comparable to previous findings (20, 21). However, there are limitations in comparing our results to prior publications due to the differences in acquisition parameters, acquisition equipment, and FreeSurfer versions that could imply confounding components (26, 27). Nevertheless, the presented data’s slope, *α*, is smaller than the previously published results. When comparing our data with a public image set, Amsterdam Open MRI Collection, AOMIC - PIOP01 (28), acquired with the same scanner and processed with the same setup, the results suggest that the slope’s variation is mainly related to age. Our supposed abnormal reported smaller *α* is probably due to the elder subjects present in the data. We cross-validated these results with the open image set AHEAD, which includes 7T MRI structural images (29). Future studies should investigate any dependency on the slope attributed to sociodemographic stats, acquisition parameters, and a bias derived from participant selection.

Nevertheless, a digest of the published results concerning the related limitations indicates that the method is robust with an acceptable variation of K, S, and I from the multiple samples (Supplementary Note 2). The results confirm the previously reported dependency of gyrification with age globally and locally (30).

### Diagnostic discrimination

To confirm that K is a better discriminator than Cortical Thickness, we estimated their optimal cut-offs and their relative accuracy, specificity, and sensibility. Lobes’ cut-offs were also determined. Our results suggest that an independent cortical morphology component, K, is of great use, even when applied to smaller ROI, in agreement with (12). We included optimal cut-off analysis for Cortical Thickness and K after removing the age effect to isolate the pathological effects in brain structure. Hereafter, K still had higher accuracy than Cortical Thickness to discriminate CTL and AD, and CTL and MCI.

Considering K is a neurodegeneration biomarker in Alzheimer’s Disease, the bi-modal shape of the curve indicates two influential groups with different levels of structural injuries, not seen by Cortical Thickness. It is possible to affirm with a visual inspection of Figure 2 that K is very sensitive to discriminate subjects in later stages of Alzheimer’s Disease or more aggressive pathological injuries. Future works should quantify if there is a connection between K and spatial distribution of brain atrophy, also an indicator of the disease later stages (31) (Supplementary Note 3). The reduced subject number limits the statistical power of including subgroups in the cut-off analysis.

When comparing CTL and MCI, the accuracy is smaller, which is expected since the MCI diagnostic includes a broad range of pathological involvement. It does not configure, in all cases, a transition to any dementia, and especially to Alzheimer’s Disease. Besides, memory loss and reduced cognitive abilities are present in healthy aging. The morphological characteristics of our MCI sample suggest that, as in the clinical aspects, the diagnostic works as an intermediate step.

Local analysis in lobes indicates K (after age correction) has better accuracy discriminating CTL and AD in the Frontal lobe, despite its very low sensibility. Higher sensibility is found on the Parietal lobe and higher specificity at the Occipital lobe. This finding confirms (21) results as the parietal lobe is affected by extended brain atrophy after the temporal lobe. The significant difference between discriminating power of K’s values with and without the age effect suggests that the parietal lobe is more affected by disease than aging. It is not possible to confirm the results from (21), which K is better discriminating Alzheimer’s Disease from healthy aging in younger individuals (Supplementary Note, Fig. 5). An extension of this work would be benefited from the increase in the number of subjects to investigate this hypothesis.

**Fig. 4.**
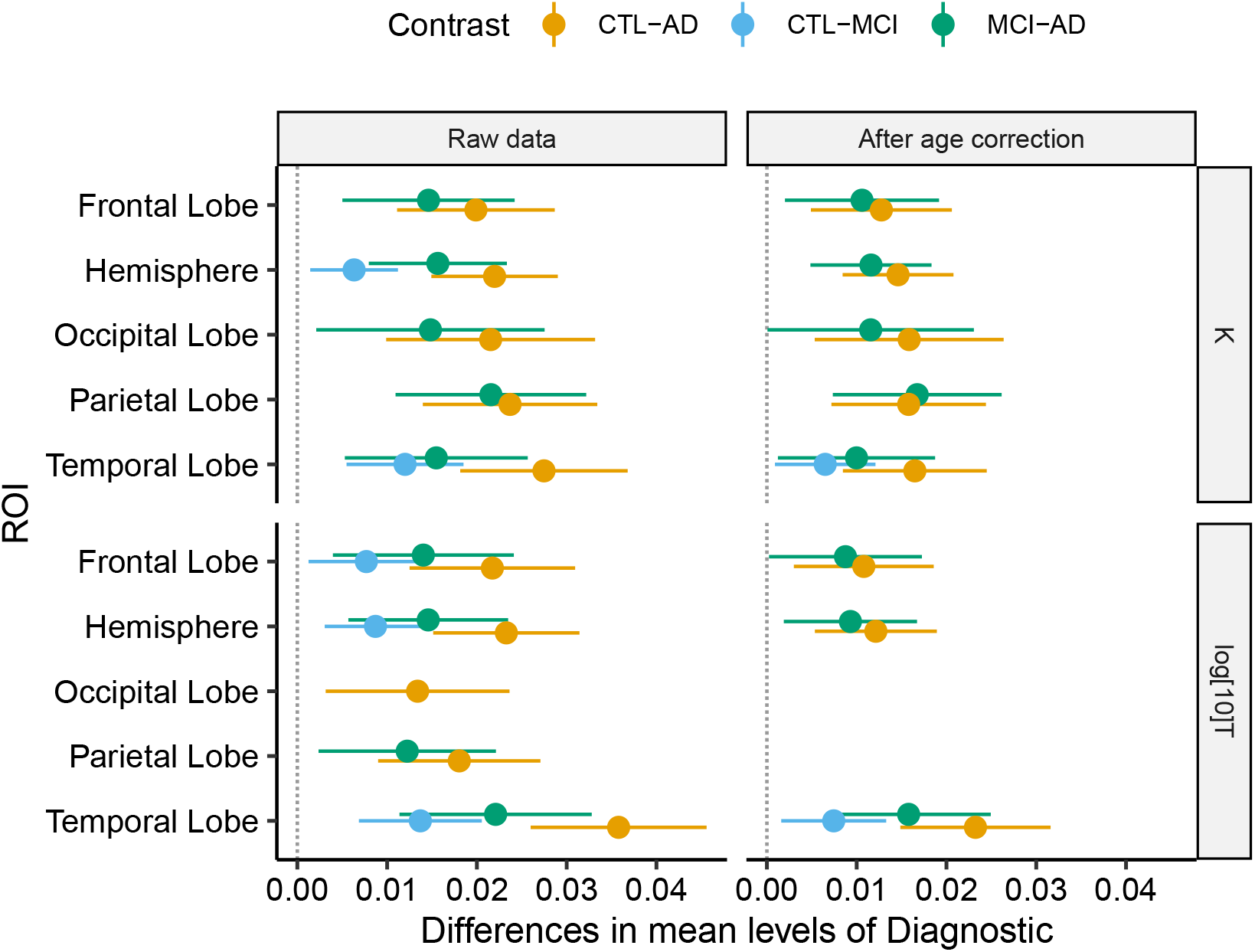
Statistically significant (p < 0.05) differences in mean levels with the 95% Confidence Interval of Diagnostics for K and log(T), with (“After age correction”) and without (“Raw data”) age correction for the hemisphere and the four lobes. Multiple corrections were applied within each morphological feature and ROI.

**Fig. 5.**
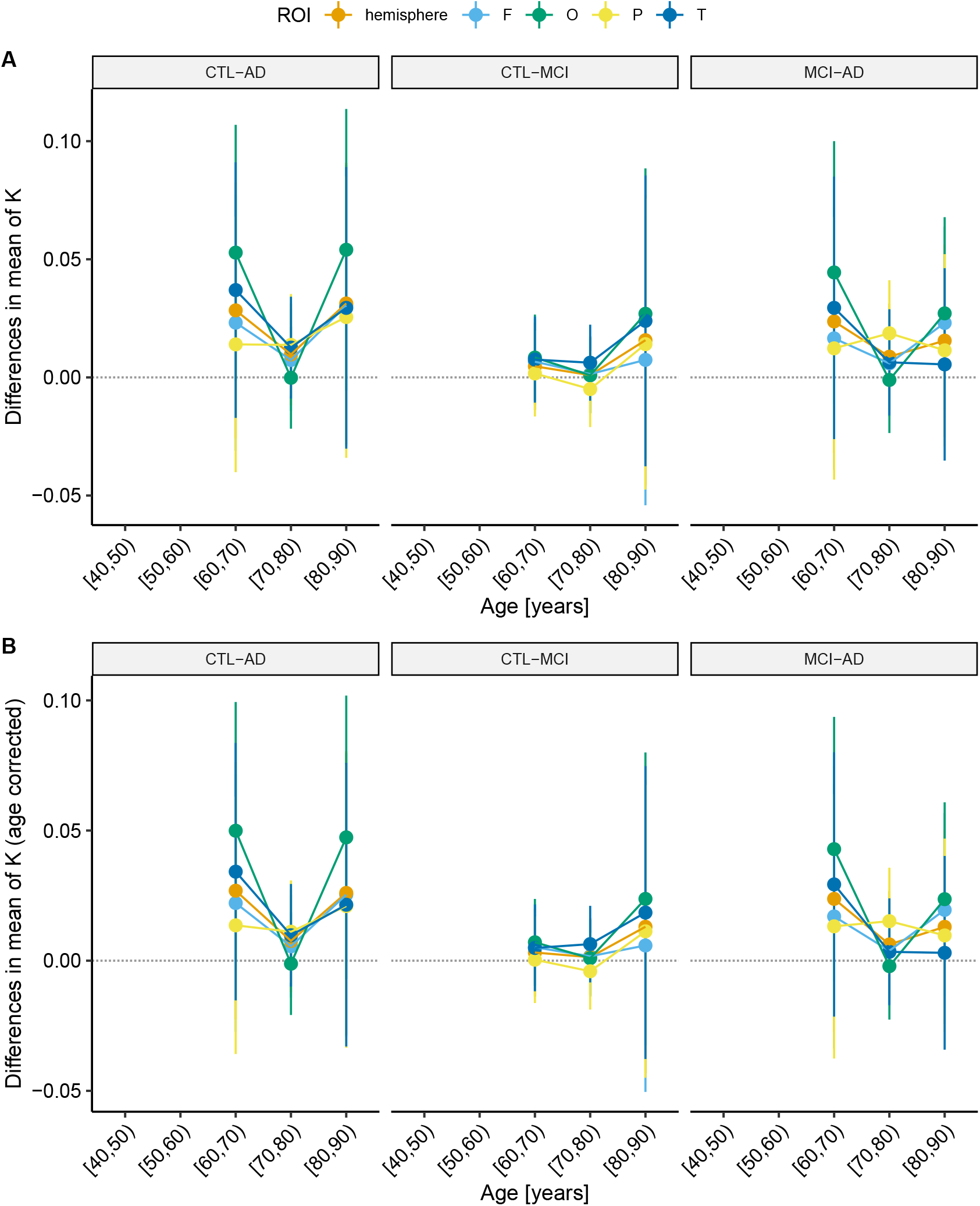
Difference of means in pairwise comparison for AD-CTL, AD-MCI, and MCI-CTL in grouped by age in decades in each ROI for (A) K and (B) K after age correction. Bars represents 95% confidence interval. There is no statistical power to infer that the difference between diagnostics is more significant in younger adults, probably influenced by the small number of observations in each data point.

### Premature and accelerated aging

As previous publications report, Alzheimer’s Disease, and healthy aging have different biological bases and onset locations of degeneration that construct both cognitive degeneration processes, from hippocampal neuronal loss (32) to the degradation of cognitive networks (33). However, AD degeneration is similar to a premature and accelerated aging process in morphological terms. Our global and local analysis corroborates this indication as K is depreciated with aging for all diagnostics. Besides, AD presents a higher unfolding rate for the four lobes. In (22), it is proposed that including S and I would improve our knowledge about brain morphology and our capability of removing aging effects. In our sample, AD visually mimics aging in K, S, and I.

Besides Cortical Thickness being one of the most studied morphological parameters for AD, further studies should consider that K represents a natural variable that translates global and local changes in brain structure and is more sensitive to less subtle changes in the disease severity. An extension of this work, one must consider i) including younger subjects to delimitate the typical values of K, S, and I; ii) increase the complexity in the discrimination model since none of the morphological parameters tested delivered results that could be used in a clinical approach; iii) verify the short and medium-term variation in cortical folding versus Cortical Thickness in a longitudinal study.

### Behavioral and biochemical correspondence

Previous studies suggested the morphological alterations in a brain with Alzheimer’s Disease are not concurrent with biochemical and behavioral alterations in AD development, as in Jack et al. (34), which suggest abnormalities in A*β*/t-Tau concentrations and brain morphology alterations precede the clinical symptoms. Moreover, one limitation of behavioral assessments is that they can include multiple cognitive domains, and most complex tasks are not mapped in a single region of the human brain. As an example, episodic memory decline can be related to the reduced number of neurons, synaptic efficiency, the concentration of neurotransmitters, and affects the prefrontal cortex, medial temporal lobe, parietal cortex, and cerebellum successively (35).

K and Cortical Thickness presented correlations at almost the same comparisons, highlighting the Cognitive Index and the episodic memory score (RAVLT A7/A5). An expected correlation was also found for t-Tau/A*β*1-42 and t-Tau/(A*β*1-42/A*β*1-40), commonly used ratios to describe Alzheimer’s Disease effects, besides the different onset time points. Therefore, regardless of the Diagnostic, the brain unfolds (measured by the decrease of variable K) with a smaller cognitive index, episodic memory score, auditory (Digit Span Backward), and visual working memory (Trail Making Test). In terms of the biochemical data analyzed, we can confirm that a less folded brain tends to have a higher concentration of t-Tau, t-Tau/A*β*1-42, and t-Tau/(A*β*1-42/A*β*1-40) ratios. We correlated K and the clinical/biochemical scores independently from the diagnostic given the non-simultaneous events.

We do not yet fully understand the contributions of deviation from biochemical and clinical typical values to the structural changes present in dementia and neurodegenerative diseases.

We can, however, provide a time-point analysis and correlate the accumulation of A*β* plaques and t-Tau tangles with reduced Cortical Thickness, and now, a less folded brain. Also, previous reports describe gyrification changes in smaller regional ROIs (36), with the association to one domain tasks and cognitive index as MMSE. Núñez investigated the association between gyrification and memory scores in AD subjects and reported significant associations for a semantic fluency test and the left insular cortex (37). In contrast, here, we use a global measure of gyrification that is theoretically motivated and shown to be correlated to multiple cognitive measurements.

## Conclusions

This manuscript intended to verify the clinical application of the proposed independent morphological components on 123 elder subjects and argues that K, the tension component, should be considered an additional structural marker to describe neurodegenerative pathologies. Our results suggest that Alzheimer’s Disease is morphologically similar to accelerated aging and distinguishable from the Mild Cognitive Impairment and Healthy Cognitive Unimpaired Control groups. It is essential to notice that K, like many morphological biomarkers, is a valuable tool to verify normal brain morphology conditions instead of determining the specific pathology that caused the brain abnormalities. Further, we demonstrated significant correlations between K and multiple behavioral tests and CSF biomarkers, which are sensitive to age correction, reinforcing that the non-concomitant process occurs during Alzheimer’s Disease development.

Finally, it is our hope that our dataset of processed 432 subjects (123 from IDOR, 208 from AOMIC PIOP01, and 101 from AHEAD) can be included in the human lifespan trajectory of K, S, and I.

A natural next step for this study would be a longitudinal study to quantify the intraindividual trajectories of K, S, and I and improvements in cortical folding sensitivity to morphological damage before AD or MCI diagnostics. Concerning the cortical folding theory proposed by Mota et al., future studies must overcome the methodological limitations of comparing samples acquired on different sites to focus on socio-economic-demographic (38) and cognitive reserve (39) and protection (40) impact on K, S, and I.

## Materials and Methods

The Alzheimer’s project sustained by IDOR is a followup study about Alzheimer’s Disease in morphological, behavioral, and biochemical aspects. The study enrolled 231 individuals from 2011 to 2018. Eligibility criteria of the project were as follows: (i) subjects had no contraindications to undergo MRI, such as presenting metal implants in the head; (ii) participants showed no signs or symptoms indicative of large-vessel cerebrovascular disease, tumoral changes, or traumatic injury affecting brain structure, as detected in clinical, cognitive and neuroimaging assessments; (iii) no severe sensorial deficits which could interfere in the application of neuropsychological tests were identified; (iv) subjects did not present major depressive disorder or any severe lifetime psychiatric disorder and (v) MRI analyses showed no significant artifacts, which could preclude the identification of brain structures. Also, subjects presenting anxiety or any other condition which interfered with his/her ability to remain still during MRI were excluded. All the participants provided written informed consent before enrollment in the study. The Hospital Copa D’Or Research Ethics Committee approved the present research under protocol number CAAE 47163715.0.0000.5249 and all images and data were anonymized after acquisition. We selected 132 subjects with structural MRI scans acquired with the same equipment and based on diagnostic criteria: Healthy subjects, Mild Cognitive Impairment, or Alzheimer’s Disease.

### Data acquisition and processing

T1-weighted MRI images (3T Philips Achieva) of the participants were acquired with the following acquisition protocol: TR/TE 7.2/3.4 ms; matrix 240×240 mm; FOV 240 mm; slice thickness 1 mm; 170 slices. The structural images were processed in FreeSurfer v6.0.0 (41) with the longitudinal pipeline (42) without manual intervention at the surfaces (43). The FreeSurfer localGI pipeline generates the external surface and calculates the local Gyrification Index (localGI) (17) for each vertex. Values of Average Cortical Thickness, Total Area, Exposed Area, and Local Gyrification Index were extracted with Cortical Folding Analysis Tool (44). We defined as ROI the whole hemisphere, frontal, temporal, occipital, and lateral lobes (based on FreeSurfer definition of lobes). The lobes’ area measurements were corrected by their integrated Gaussian Curvature, removing the partition size effect and enabling a direct comparison between lobes and hemisphere cortical folding (21). Due to processing errors, eleven subjects were excluded during the FreeSurfer processing or data extraction steps. The final number of subjects included in this report is 123 (77 CTL, 33 MCI, and 13 AD).

A team of physicians, psychologists, and speech therapists handled the Clinical behavior and the biochemical assessment as described previously (45). The tests included Digit Span Backwards, RAVLT A7, RAVLT A5, and Trail Making Test (TMT). The Cognitive Index is calculated as a global cognitive function (composed of TMT and RAVLT) weighted for age intervals of 10 years. For a subset of our sample (described in Supplementary Note, Table 3), we included the following biochemical biomarkers from the Cerebrospinal Fluid (CSF): Lipoxin, A*β*1-42, A*β*1-40, and t-Tau.

**Table 3.** Summary of each sociodemographic, morphological, behavioral, and biochemical variable. Mean values *±* standard deviation (number of subjects).

| Variable | CTL (N = 77) | MCI (N = 33) | AD (N = 13) |
| --- | --- | --- | --- |
| Age [years] | 65.87 ± 8.42 | 72.25 ± 4.62 | 77.08 ± 6.14 |
| Education [years] | 15.22 ± 2.20 | 13.18 ± 2.41 | 12.62 ± 2.98 |
| Female, N (%) | 53 (69%) | 19 (51%) | 8 (62%) |
| Morphometrical |  |  |  |
| Cortical Thickness [mm] | 2.51 ± 0.10 | 2.46 ± 0.09 | 2.38 ± 0.08 |
| Total Area [mm<sup>2</sup>] | 98420.20 ± 7836.81 | 97382.63 ± 8529.96 | 95109.22 ± 9251.56 |
| Exposed Area [mm<sup>2</sup>] | 37478.26 ± 2395.69 | 37306.01 ± 2795.69 | 37160.08 ± 2962.07 |
| k | 0.30 ± 0.01 | 0.29 ± 0.01 | 0.28 ± 0.01 |
| K ($\log_{10} k$) | -0.52 ± 0.01 | -0.53 ± 0.01 | -0.55 ± 0.02 |
| S | 9.1 ± 0.01 | 9.2 ± 0.12 | 9.2 ± 0.13 |
| I [mm<sup>6</sup>] | 10 ± 0.06 | 10 ± 0.07 | 10 ± 0.07 |
| Behavioral |  |  |  |
| Cognitive Index | 0.21 ± 0.64 (77) | -1.48 ± 1.27 (33) | -3.35 ± 1.46 (13) |
| RAVLT A7/A5 | 0.82 ± 0.18 (77) | 0.54 ± 0.30 (33) | 0.24 ± 0.31 (13) |
| TMT B-A | 58.67 ± 47.57 (77) | 139.32 ± 110.78 (33) | 226.69 ± 129.06 (13) |
| Digit Span Backward | 5.84 ± 1.74 (77) | 4.70 ± 1.59 (33) | 3.77 ± 1.37 (13) |
| Biochemical |  |  |  |
| Lipoxin [pg/mL] | 127.15 ± 61.04 (28) | 118.35 ± 47.33 (14) | 79.10 ± 70.87 (6) |
| Aβ1-40 [pg/mL] | 4192.04 ± 1900.51 (29) | 4935.29 ± 2783.09 (14) | 5664.22 ± 1603.33 (6) |
| Aβ1-42 [pg/mL] | 533.92 ± 240.92 (29) | 433.47 ± 299.05 (14) | 279.71 ± 57.75 (6) |
| t-Tau [pg/mL] | 354.87 ± 193.47 (29) | 449.59 ± 187.44 (14) | 632.00 ± 268.36 (6) |

All statistics were analyzed with R v4.1.0 and has the code available at GitHub (46). Multiple comparisons of means were made with ANOVA, post hoc evaluations with Tukey multiple comparisons of means (which presents p-value corrected for multiple comparisons, “p adj”), correlation tests with Pearson’s r and Cohen’s d Effect Size. Correlation’s p-value was corrected for multiple comparisons (Bonferroni) within the clinical assessment and morphological parameters. K, S, and I were normalized to unity vector when necessary. Cut-offs were determined, maximizing the sensitivity and specificity’s sum with bootstrapping to estimate the cut point variability (bootstrap number = 1000). The statistical significance threshold was *α* = 0.05.

## Data Availability

Data acquired at IDOR does not have clearance for public sharing patients information that could lead to identification due to local Ethics Committee approval restrictions. Processed structural MRI images from the AHEAD and AOMIC datasets are available at open repository.

https://doi.org/10.5281/zenodo.5750618

## ACKNOWLEDGEMENTS

This work was funded by the Research Support Foundation of the State of Rio de Janeiro (FAPERJ), National Council for Scientific and Technological Development (CNPq), and intramural grants from Instituto D’Or de Pesquisa e Ensino (IDOR). Bruno Mota is supported by Fundação Serrapilheira Institute (grant Serra-1709-16981) and CNPq (PQ 2017 312837/2017-8). We are thankful to the research team and volunteers for participating in this research project.

## Supplementary Note 1: Extended Results

Extended results with supplementary Tables and Figures. Table 3 describes diagnostic groups in Sociodemographic, Morphometrical, Behavioral and Biochemical information. Figure 4 compares post hoc difference of means for K and Average Cortical Thickness in all ROIs with and without age correction. Figure 5 compares post hoc difference of means comparing diagnostic means of K and K (age-corrected) at each age decade to explore whether the difference would be related to age. Table 4 presents the extended results K (raw data) optimal cut-off analysis for discriminating Alzheimer’s Disease, Mild Cognitive Impairment, and Cognitive Unimpaired Controls. Table 5 presents the extended results for the correlation analysis within K or Average Cortical Thickness and behavioral and biochemical biomarkers of Alzheimer’s Disease.

**Table 4.** Optimal cut-off (maximum sensitivity + specificity) for K at each ROI (hemispheres and lobes). Cut-off, accuracy, sensibility, and specificity for discriminating pairwise diagnostic groups.

| ROI | CTL-AD |  |  |  | CTL-MCI |  |  |  |
| --- | --- | --- | --- | --- | --- | --- | --- | --- |
|  | Cut-off | Acc | Sens | Spec | Cut-off | Acc | Sens | Spec |
| Hemi | -0.54 | 0.82 | 0.58 | 0.86 | -0.53 | 0.60 | 0.55 | 0.63 |
| Frontal | -0.55 | 0.69 | 0.65 | 0.69 | -0.54 | 0.55 | 0.69 | 0.49 |
| Occipital | -0.50 | 0.79 | 0.58 | 0.58 | -0.48 | 0.51 | 0.51 | 0.51 |
| Parietal | -0.52 | 0.75 | 0.77 | 0.75 | -0.50 | 0.46 | 0.57 | 0.42 |
| Temporal | -0.51 | 0.69 | 0.73 | 0.68 | -0.51 | 0.63 | 0.58 | 0.66 |

**Table 5.**
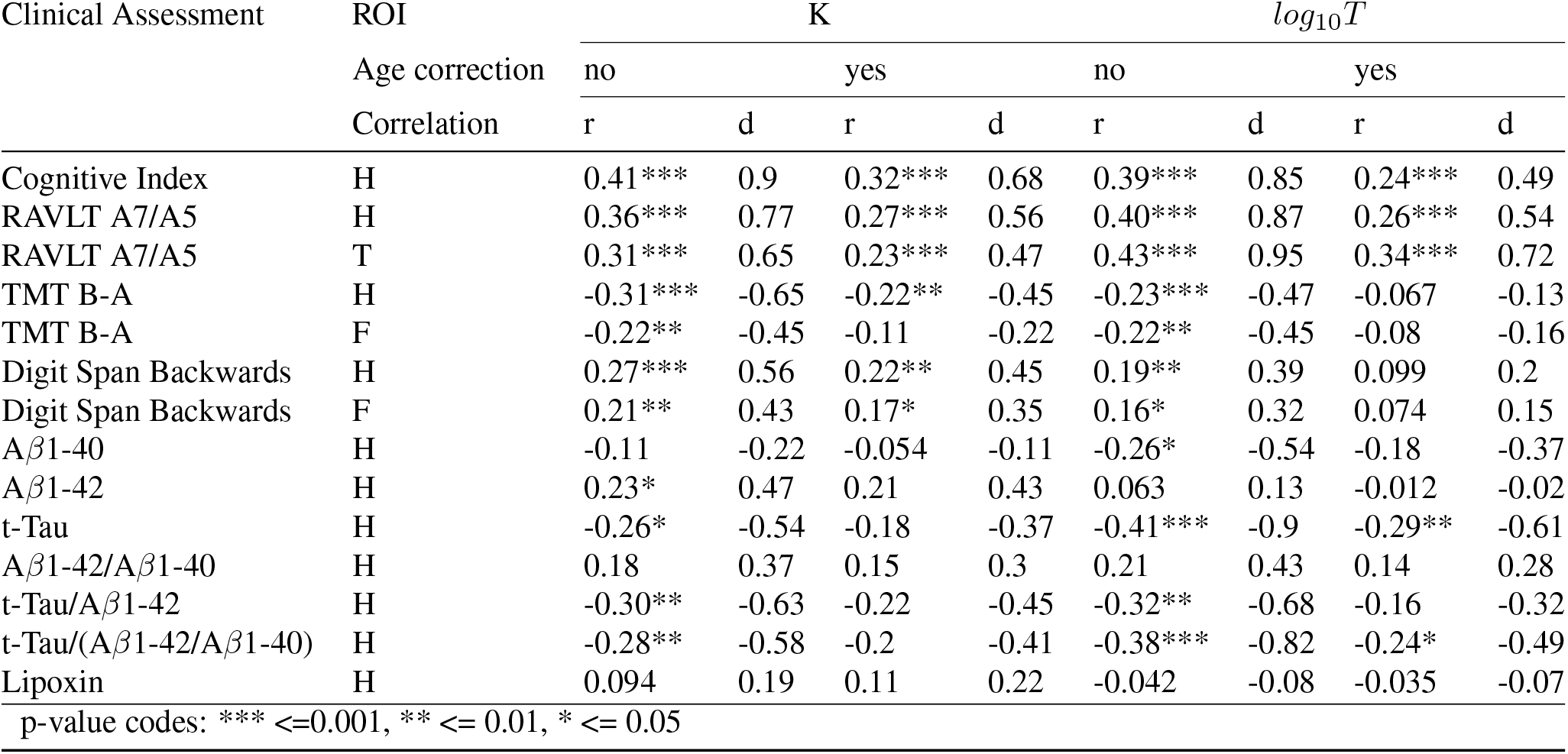
Extended table with Pearson’s r correlation results with Cohen’s d effect size for behavioral assessments and morphological parameters, K and Average Cortical Thickness, with and without age correction. P-value was corrected (Bonferroni) for multiple comparisons within Clinical Assessment and morphological measurement. ROI codes: H - Hemisphere, F - Frontal Lobe, O - Occipital Lobe, P - Parietal Lobe and T - Temporal Lobe.

## Supplementary Note 2: Slope interpretation

One unexpected aspect of the reported data in the manuscript is that the data presents a smaller cortical folding model slope (Figure 6) than the previously published results (20). Considering that multiple acquisition and processing sites increase the variability in brain morphology studies (26, 27, 47–49), the slope analysis should be handled by reducing the possible methodological confounding variables.

**Fig. 6.**
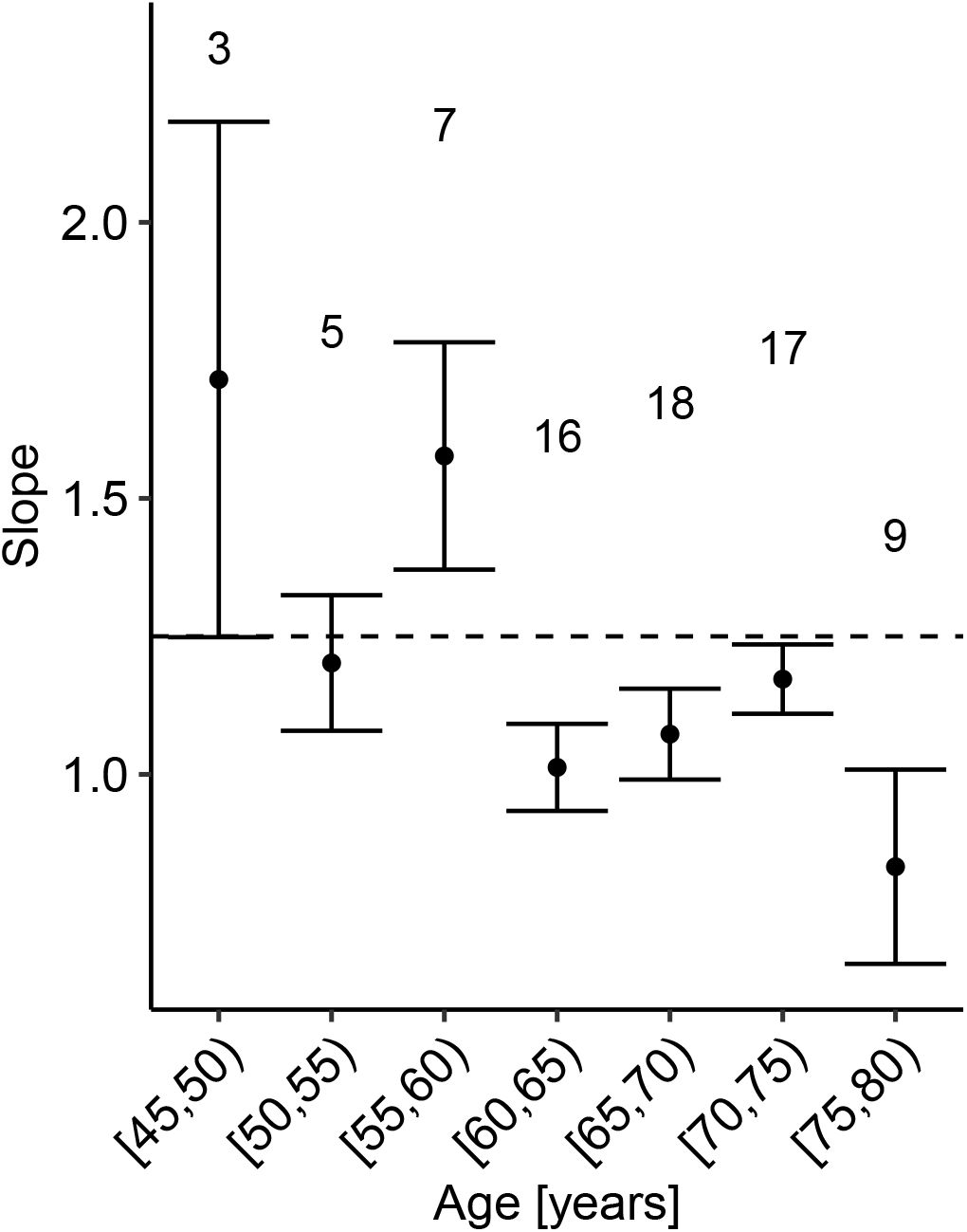
Age effect in cortical gyrification. Cortical folding model slope *α* within 5 years, only for Healthy Cognitive Unimpaired Controls. Above each point, we display the number of subjects in each regression. Bars represent the standard deviation for the respective age interval regression. Points with one subject were excluded due to the lack of statistical significance.

We compared the data of this manuscript with the Amsterdam Open MRI Collection (AOMIC) PIOP01 dataset. MRI T1-weighted images were acquired at the same MRI equipment, Philips Achieva 3 T. Moreover, we cross-validated the results with the Amsterdam Ultra-High Field Adult Lifespan database (AHEAD), a 7 T available set of MR images.

AOMIC PIOP01 and AHEAD were processed with the FreeSurfer v6.0 (41) standard pipeline, the localGI FreeSurfer pipeline, and the Cortical Folding Analysis Tool (44). AOMIC PIOP01 dataset (28) contains 216 healthy younger subjects close to 25 years old, the age used to correct our data. Seven subjects were excluded for missing age information and one due to error in processing and extraction. AHEAD dataset (29) contains 105 healthy subjects from 18 to 80 years old. From those, four subjects were excluded due to errors in processing or uncorrected surface. Processed data from both datasets are available (50). We included S and I values for the datasets published by Wang et al. (20) (Table 6).

**Table 6.**
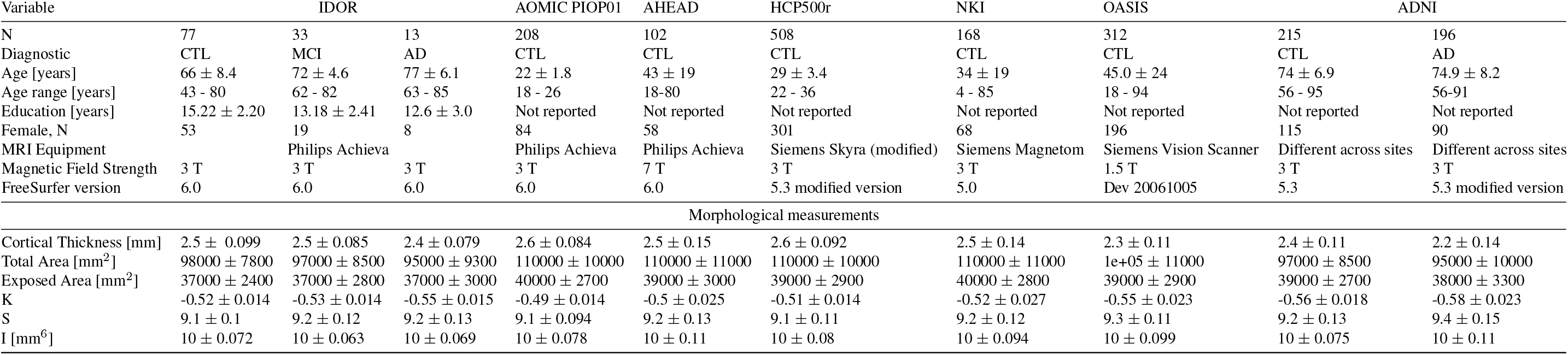
Description of the datasets included in this report, summarizing each morphological, cognitive, and biochemical test. Values are mean values *±* standard deviation. Diagnostic code: CTL for Controls, MCI for Mild Cognitive Impairment, and AD for Alzheimer’s Disease.

The IDOR slope is smaller than HCP500r, AHEAD, AOMIC, NKI, and OASIS and comparable to ADNI-Control and ADNI-AD, both samples with exclusively elder subjects (Fig. 7). To confirm the hypothesis of slope dependency on age without the methodological confounding variables, we verified the slope behavior in AOMIC and IDOR-Control samples through Age (Fig. 8 - A). There is a tendency of a reduced slope with the increase of age, and this tendency is similar to the negative gradient found in cortical thickness and K (Fig. 8 – B and C). These results are validated when compared to the AHEAD subjects (Fig. 9). After a certain age, between 40 to 60 years, the slope escapes the model’s expectations, which could be explained by the nonhomogeneity in aging effects on brain structure. Aging occurs with different onsets, locations, and scales (from a single cerebral gyrus to a whole lobe).

**Fig. 7.**
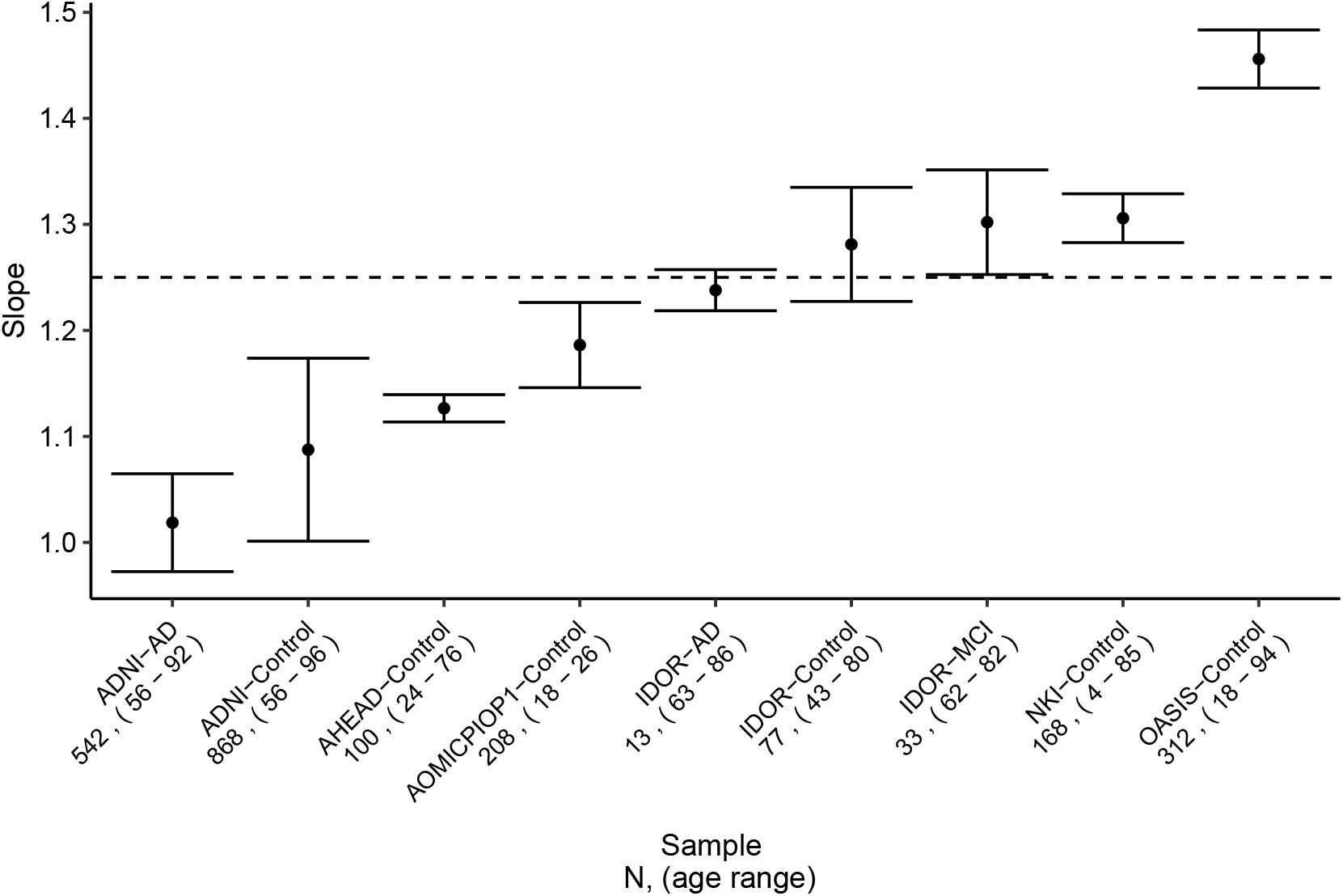
Slope for each sample with the number of subjects included and the age range. The traced line is for 1.25, the theoretical value of the slope *α*. Bars represents the standard deviation.

**Fig. 8.**
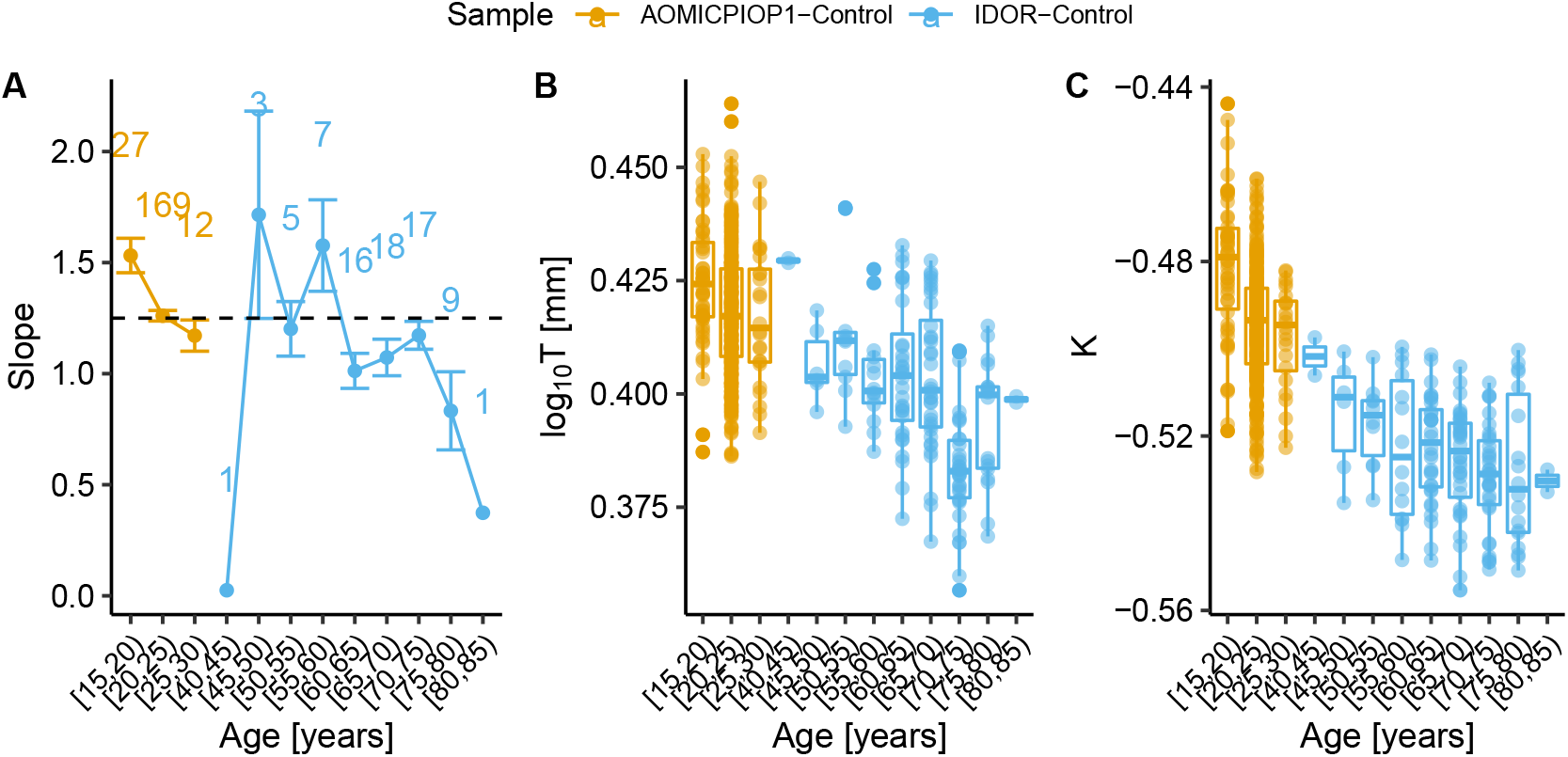
Plots comparing morphological variables behavior with Age. Red is for AOMICPIOP01-Control and blue for IDOR-Control. The x-axis is modified to hide the age interval from 30 to 40 years old that has none subjects, two data points with only one subject, 40-45 and 80-85. The total number of subjects included are 284. (A) Slope for each Age interval group and each sample. The number on top of the point indicates the number of subjects included in the linear regression, and each subject contributes with two data points, one for each hemisphere. The traced line is for 1.25, the theoretical value of the slope. Bars represents the standard deviation. (B) Log10(T) distribution for each Age interval group and each sample. Bars represents the 95% Confidence interval. (C) K distribution for each Age interval group and each sample. Bars represents the 95% Confidence interval.

**Fig. 9.**
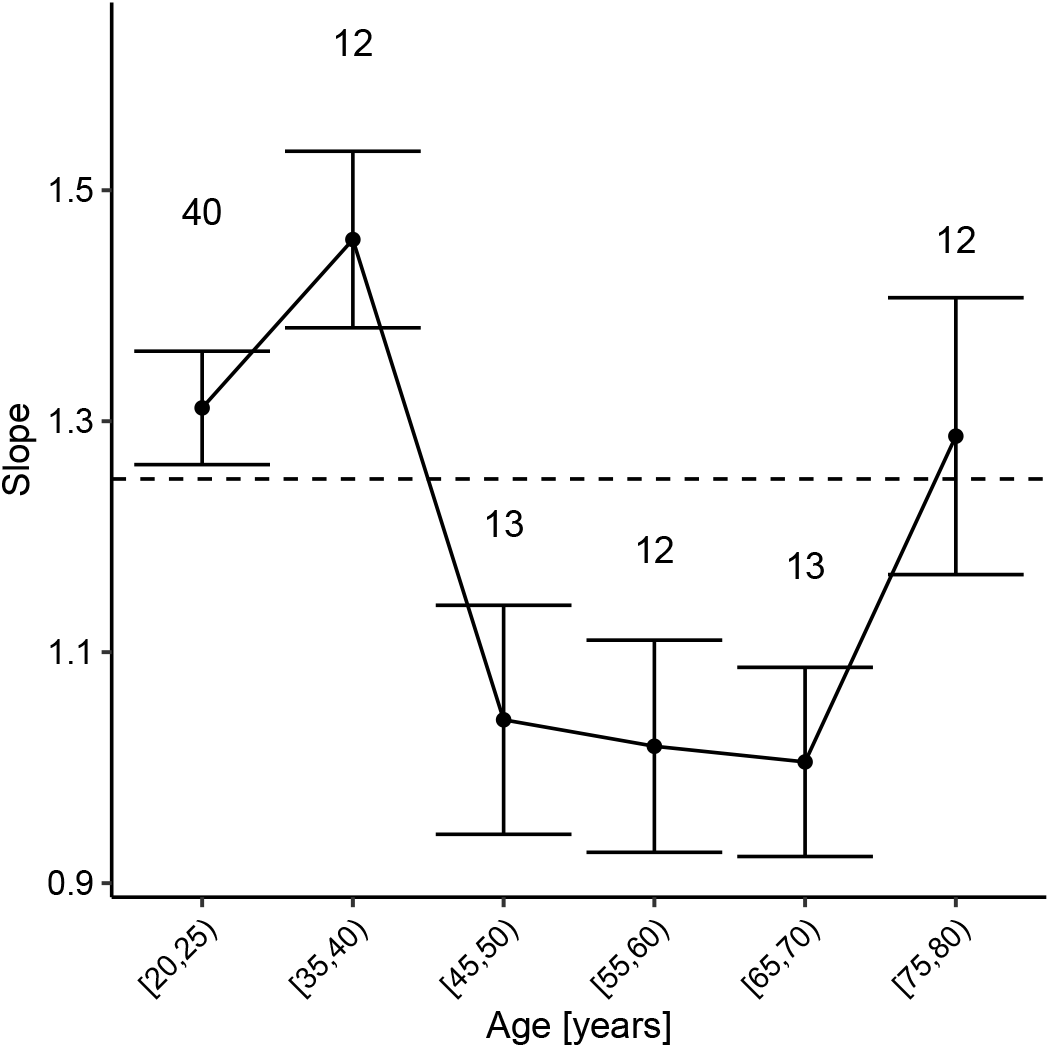
Slope for each Age interval group in AHEAD-Control. The number on top of the point indicates the number of subjects included in the linear regression, and each subject contributes with two data points, one for each hemisphere. The traced line is for 1.25, the theoretical value of the slope *α*. Bars represents the standard deviation.

Future works with data from a unique site and protocol could evaluate the effect of socioeconomic status and prospect the biological meaning of the slope.

This manuscript also contributes to sketch the trajectory across the human lifespan for K, S, and I. The fitting of Control data points through age demonstrates an acceptable variance considering the methodological limitations and intraspecies biological diversity (Fig 10). The age dependency in K (Pearson’s r = -0.77, p < 0.0001) and I (Pearson’s r = -0.62, p < 0.0001 is confirmed. The shape factor has the most intersubject variability possibly, but still significant correlation (Pearson’s r = 0.21, p < 0.0001.

**Fig. 10.**
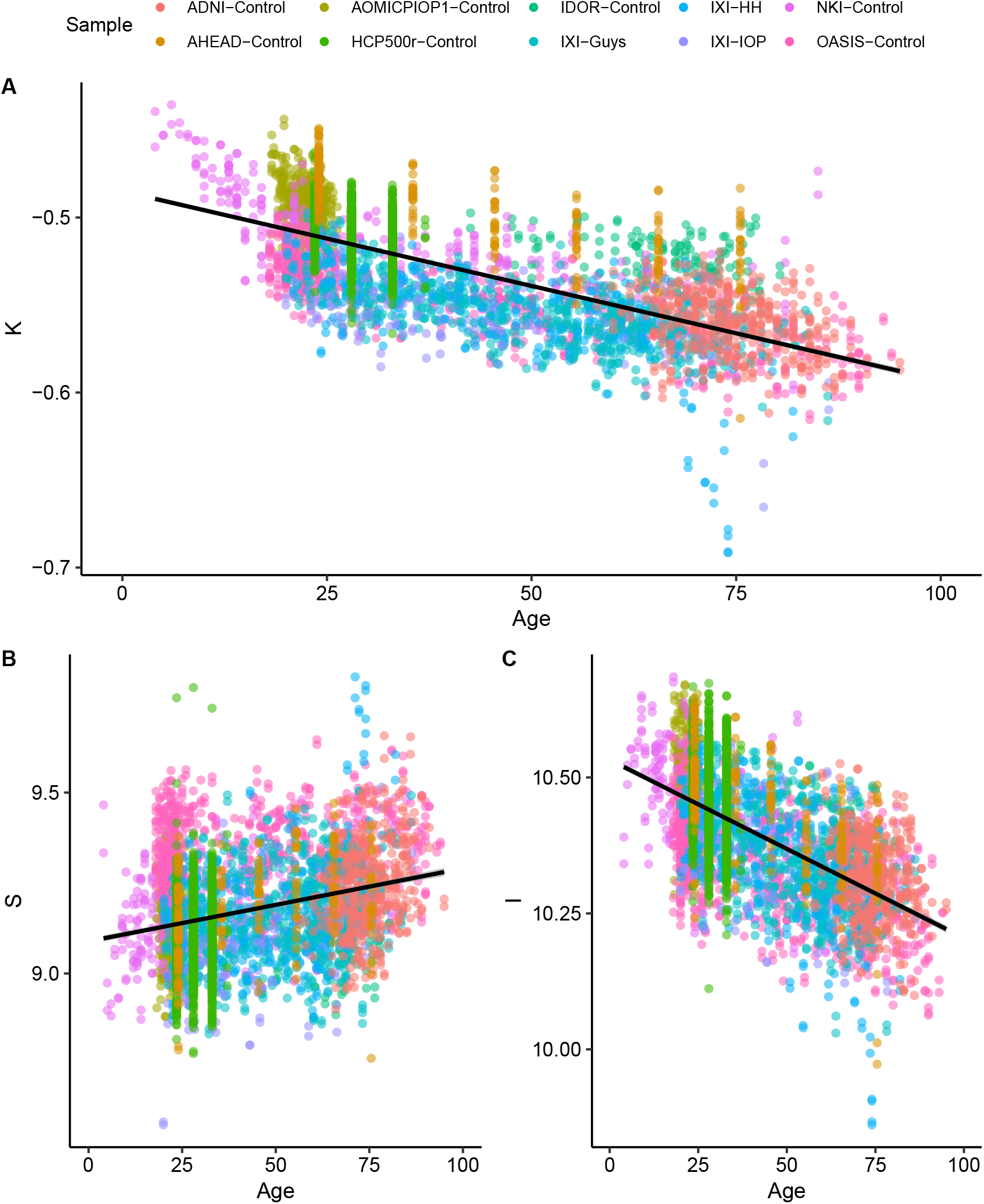
Distribution of subjects for every Healthy Control subject across the independent morphological component through age. For HCP500r and AHEAD, the mean age of the interval was considered, since ages are determined by an interval of years, instead of one value for each subject. The solid line represents a linear regression applied for all data with the 95% confidence interval. (A) K, the tensor component, Pearson’s r = -0.77, p < 0.0001 (B) S, the shape component, Pearson’s r = 0.21, p < 0.0001, and (C) I, the volume component, Pearson’s r = -0.62, p < 0.0001.

## Supplementary Note 3: Bimodal distribution of K in Alzheimer’s Disease and optimal cut-off analysis

As commented in the Manuscript Discussion, the bimodal distribution of K is very prominent for the AD subjects. However, the reduced number of subjects limits our statistical and inference power. Nevertheless, the bimodal distribution communicates that we have two groups divided by the intensity of whole-brain structural injuries in this sample. Lower values of K are related to healthy aging and Alzheimer’s Disease as proved here and in (20). Thereby, based on Alzheimer’s Disease spatial evolution at the human brain (31), one could hypothesize that the worst the structural damage, the spreader the damage would be, and the lower K values would be found. Based on this hypothesis, we estimated the minimum value of the valley between peaks with a simple mathematical procedure (value of K were *f*′(*K*) = 0. The valley is placed at K = -0.5432, with 11 hemispheres with K >-0.54321 and 15 hemispheres with K < -0.5432, with a total of 26 hemispheres for the 13 subjects as expected. Those hemispheres were divided into two groups, and then we visually evaluated the density plots regarding the K for the lobes of those hemispheres Figure 11. Here, we can verify that hemispheres with lower K and consequently more significant distance between AD and CTL/MCI peaks (meaning more considerable discriminating power) had the same pattern at the lobes.

**Fig. 11.**
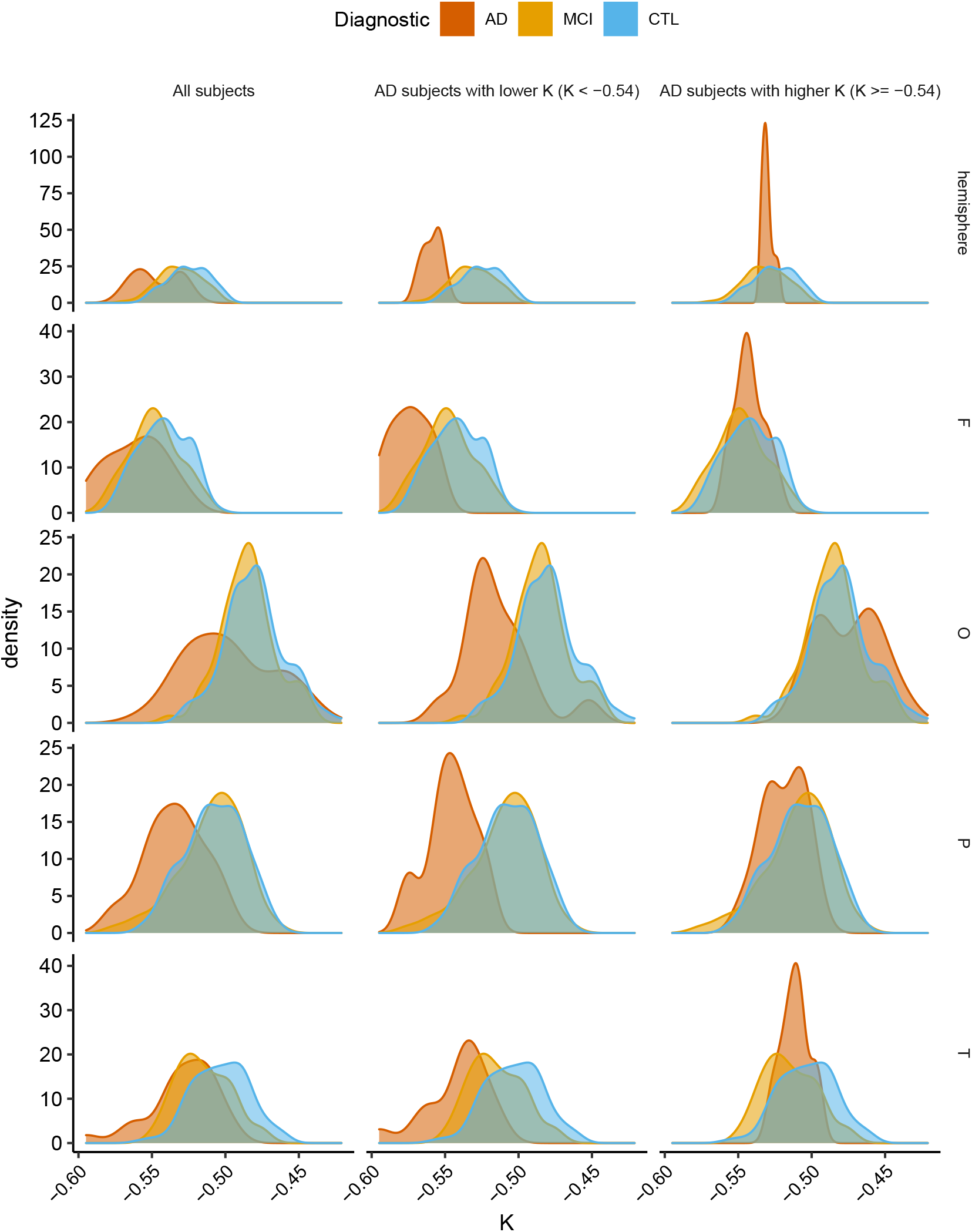
K density plots across hemisphere and lobes. AD as red, MCI as green, and CTL as blue. In the first column, all AD subjects were included, along with all MCI and CTL subjects. The second column display results only for AD subjects with hemispherical K < -0.54, and MCI and CTL subjects. Finally, all AD subjects with hemispherical K > -0.54 or K = -0.54 with MCI and CTL subjects

We confirmed the increase of the discriminating power by including only AD subjects with K < -0.5432187 in the optimal cut-off analysis; The accuracy in discriminating AD and CTL has an impressive increase to 0.96, while the sensibility and specificity rise to 0.93 and 0.97, respectively (Figure 12). The optimal cut-off in this analysis is -0.55. As expected, the discriminating power of diagnostics using the Cortical Thickness as a biomarker also increases. However, it does not reach K discriminating power levels.

**Fig. 12.**
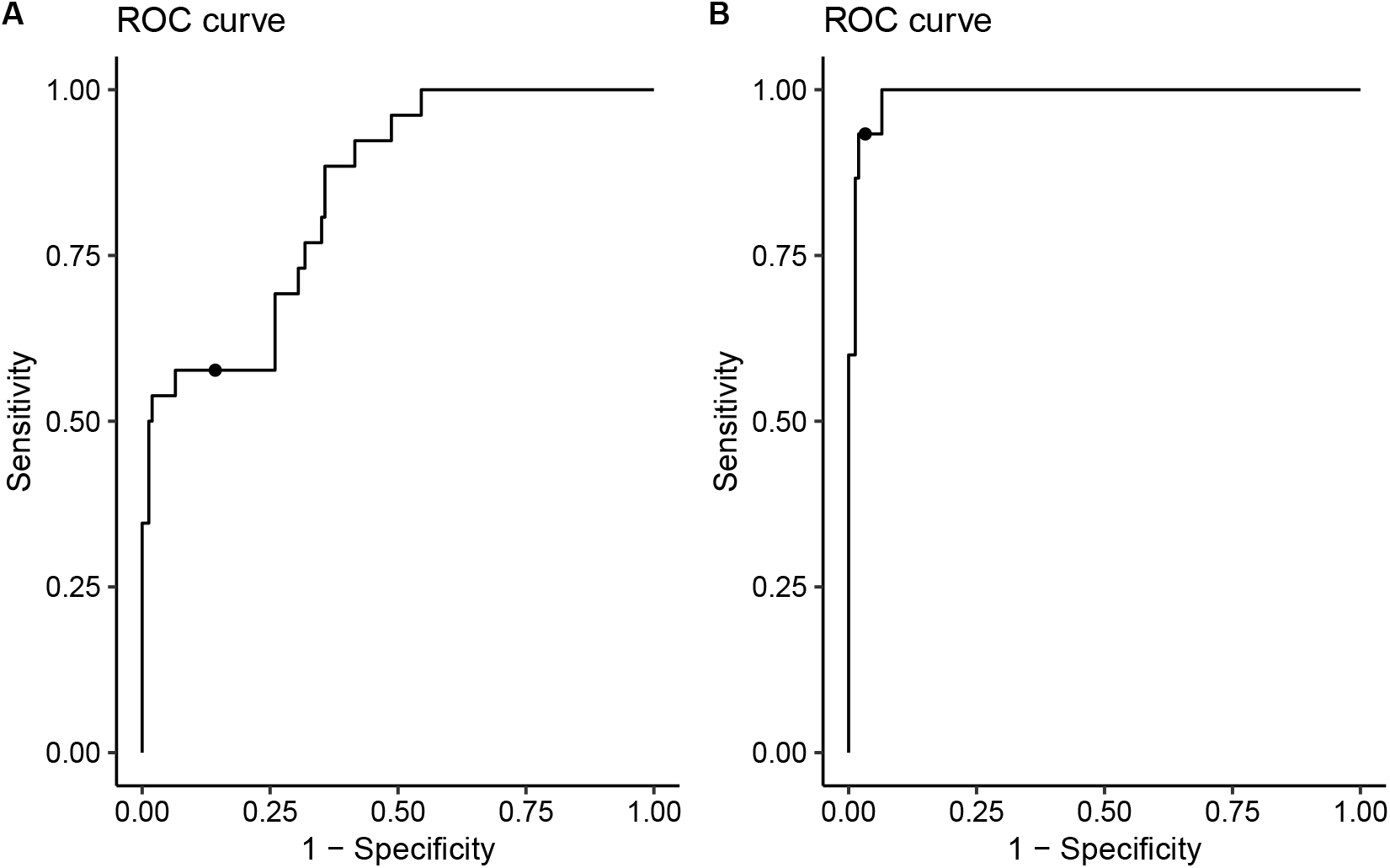
ROC curves derived from the optimal cut-off analysis of K (hemisphere as ROI) to discriminate CTL and AD subjects. (A) All AD subjects included and optimal cut-off = -0.54. (B) AD subjects included if hemispherical K < -0.54, optimal cut-off = -0.55.

